# ESTIMATING COVID-19 INFECTIONS IN HOSPITAL WORKERS IN THE UNITED STATES

**DOI:** 10.1101/2020.04.06.20055988

**Authors:** Junaid A. Razzak, Junaid A. Bhatti, Ramzan Tahir, Omrana Pasha-Razzak

**Affiliations:** Johns Hopkins University School of Medicine, USA, and the Aga Khan University College of Medicine, Pakistan; Independent Researcher, Ajax, ON, Canada; Apotex Inc., Toronto, ON

**Author notes:** **CORRESPONDING AUTHOR** Junaid A. Razzak, MD, PhD, FACEP, Johns Hopkins University School of Medicine, 5801 Smith Avenue, Suite 320, Baltimore, Maryland 21209.

**Keywords:** Corona virus, COVID-19, hospital workers, Personal Protective Equipment (PPE), SARS-COV2

## Abstract

**Objective:** We estimated that how many hospital workers in the United States (US) might get infected or die in the COVID-19 pandemic. We also estimated the impact of personal protective equipment (PPE) and age restrictions on these estimates.

**Methods:** Our secondary analyses estimated hospital worker infections in the US based on health worker infection and death rates per 100 deaths from COVID-19 in Hubei and Italy. We used Monte Carlo simulations to compute point estimates with 95% confidence intervals for hospital worker infections in the US based on the two scenarios. We computed potential decrease in infections if the PPE were available only to those involved in direct care of COVID-19 patients (∼ 30%) and if workers aged ≥ 60 years are restricted from patient care. Estimates were adjusted for hospital workers per bed in the US compared to China and Italy.

**Results:** The hospital worker infections per 100 deaths were 108.2 in Hubei and 94.1 in Italy. Based on Hubei scenario, we estimated that about 53,640 US hospital workers (95% CI: 43,160 to 62,251) might get infected from COVID-19. The Italian scenario suggested 53,097 US hospital worker (95% CI: 37,133 to 69,003) might get infected during the pandemic. Availability of PPE to high-risk workers could reduce counts to 28,100 (95% CI: 23,048 to 33,242) considering Hubei and to 28,354 (95% CI: 19,829 to 36,848) considering Italy. Restricting hospital workers aged ≥ 60 years from direct patient care reduced counts to 1,985 (95% CI: 1,627 to 2,347) considering Hubei and to 2,002 (95% CI: 1,400 to 2,602) considering the Italian scenario.

**Conclusion:** We estimated significant burden of illness due to COVID-19 if no strategies are adopted. Making PPE available to all hospital workers and reducing exposure of hospital workers above the age of 60 could have significant reductions in hospital worker infections.

**VISUAL ABSTRACT:** Figure 1.
Estimated number of COVID-19 related infections among healthcare workers in the United States based on Hubei and Italian scenarios

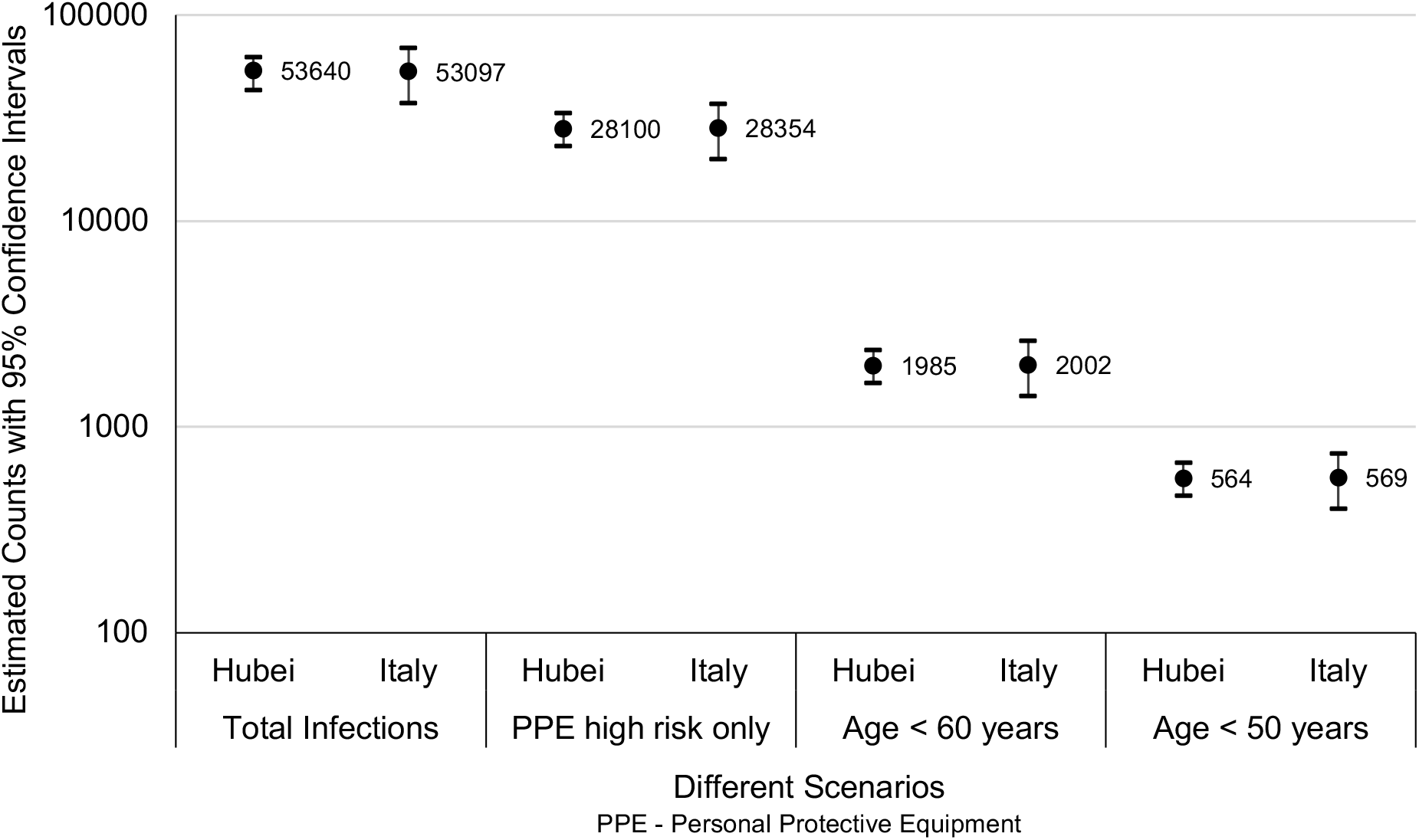

## INTRODUCTION

The COVID-19 pandemic has resulted in significant loss of life and major disruption of social and economic structures across the globe.^1^ By March 27th, 2020, the total number of people infected exceeded 595,000 with over 27,000 deaths.^2^ Even robust healthcare systems are challenged severely by the numbers of patients and the severity of the illness.^3^

One of the major concerns in managing this outbreak is the safety of healthcare workers especially those working in the hospital settings.^4^ The city of Wuhan in China has seen over 3,000 healthcare workers being infected while in Italy, where a major epidemic is ongoing has reported 7,145 healthcare worker infections including 51 physician deaths by March 27, 2020.^5^ Ongoing shortages of personal protective equipment (PPE) have resulted in heightened anxiety and in some cases refusal of care by the healthcare providers.^6^ In the face of restrictions to PPE availability and usage, as well as case reports of deaths of doctors and nurses, hospital workers are worried about their health, their ability to keep working and possible mortality, as well as the risk to their families.^7^ These concerns are not unfounded as the risk of infection appears inordinately high, with 20% of responding hospital workers in Italy becoming infected.^3^ While hospital workers without the requisite PPE continue to see patients in many settings, an increasing number of hospital worker infections or increasing reluctance to provide care remains one of the major risks to the global response.^4^

There is currently no published evidence on the expected number of infected hospital workers in the US or projections for other countries around the world.^8^ The available data show that the risk of COVID-19 infection amongst hospital-based workers is directly proportional to the number of patients in the healthcare system.^5,9,10^ This relationship is modulated by other factors, including adequate supply and optimal usage of PPE.^4,6,8,10^ The shortage of PPE has created circumstances where its use is prioritized for hospital workers that are highly exposed to COVID-19 patients, such as intensive care or emergency care workers.^9,11,12^ Illness outcomes, including mortality, are further influenced by the characteristics of the hospital worker population, including age structure and prevalence of other risk factors, such as occurrence of comorbid conditions, which could have a direct impact on risk of disease severity and number of deaths.^1,5,13-15^

We estimated the number of hospital workers in the US at risk of contracting the infection or dying due to COVID-19. We estimated COVID-19-related counts for infections and deaths if perfect PPE conditions are available to high risk hospital workers. We also evaluated the impact of curtailing hospital-based direct patient care in workers above a certain age on estimated counts for infections and deaths.

## METHODS

### Study design and settings

Our secondary analyses estimated hospital worker infections in the US based on health worker infections and deaths per 100 deaths from COVID-19 in Hubei and Italy normalized for hospital workers per beds in the US.

### Data Sources

We extracted data for COVID-19 infections from Hubei, China and Italy. We also extracted data from the two other jurisdictions, the Wuhan city (located in Hubei province) and the country of South Korea for initial comparisons. China’s Hubei province, the initial site of the epidemic is a landlocked province with a population of more than fifty-eight million people, with the city of Wuhan home to almost eleven million of them. Wuhan is a transport hub and major rail interchange in China and was at the center of the initial outbreak, with the first case reported on December 1, 2019. While cases spread to the rest of the province and, during the annual Chinese New Year migration, to other provinces; the city remained the epicenter of the outbreak in China, with more than 60% of all cases in the country. With widespread quarantine measures, the outbreak in China appears to be largely controlled at this time and no new cases have been reported since March 19, 2020. Italy, which surpassed China in the number of deaths due to COVID-19 and it is still in the middle of the epidemic.^5^ The outbreak here has centered around the city of Bergamo in the province of Lombardy. Approximately, half of the cases in the country are in Lombardy and the numbers of deaths continue to rise at an unprecedented rate.^16^ South Korea, a nation of 51 million people, is recognized as an exemplar in controlling the spread of the disease and reporting one of the lowest mortality rates in the world.^17-19^ We used the publicly available data covering all cases from the beginning of the epidemic till March 19, 2020 for China and South Korea and till March 27 for Italy.

#### Coronavirus COVID-19 Global Cases by the Center for Systems Science and Engineering (CSSE) at Johns Hopkins University^2^

Initially based on DXY, an online platform run by members of the Chinese medical community, which aggregates local media and government reports to provide COVID-19 cumulative case totals in near real-time at the province level in China and country level otherwise.^20^ Every 15 minutes, the cumulative case counts are updated from DXY for all provinces in China and affected countries and regions. Additionally, for countries and regions outside mainland China (including Hong Kong, Macau and Taiwan), other sources include twitter feeds, online news services, and direct communication sent through the dashboard, that are confirmed with regional and local health departments, including the China CDC (CCDC), Hong Kong Department of Health, Macau Government, Taiwan CDC, European CDC (ECDC), the World Health Organization (WHO), as well as city and state level health authorities.^2^ For city level case reports in the U.S., Australia, and Canada reporting relies on the US CDC, Government of Canada, Australia Government Department of Health and various state or territory health authorities. All manual updates are coordinated by a team at JHU. The data are stratified by country and include total confirmed cases; daily new cases; total confirmed deaths; daily new deaths and recovered cases.^2^ Information is not available on tests or breakdown of confirmed cases.

#### Official government publications

We used the daily report published by the Italian Ministry of Health (English version) to obtain the overall number of COVID-19 infections, deaths amongst those patients as well as the number of health worker infections.^5^ We also obtained the percentage of patients who were severely or critically ill from the report. For US data, we used the most up to date official press release from the city and state departments of New York,^21^ Massachusetts,^22^ and Florida.^23^

We used the Organization of Economic Cooperation and Development data for 2017 for numbers of physicians, nurses, and hospital beds in China, Italy, and the United States.^24,25^

### Study measures

From all three sites, we attempted to extract data regarding the total number of fatalities, the number of health workers found to be infected with COVID-19 and health worker mortality. Number of fatalities – defined as the number of cases reported as a case of suspected, probable or confirmed COVID-19 that died as a result of the disease or its complications.

Healthcare workers infected –For the integrated surveillance of COVID-19 in Italy, laboratory confirmed infection with COVID-19 amongst people who meet the definition of COVID-19. A similar definition was adopted by the Chinese Center for Disease Control and Prevention, including providers of medical treatment and care services

### Statistical Analyses

We estimated the hospital worker infections and deaths in the US in systematic stepwise computations based on observed statistics from Hubei and Italy. Firstly, we computed the COVID-19 deaths per million population. We then calculated the number of admissions based on the proportion of severe and critical cases. Based on reported health worker infections in Hubei and Italy, we estimated the rates of healthcare worker infections per 1,000 admissions and per 100 deaths.

We then computed the expected deaths in the US based on COVID-19 deaths per million population in various jurisdictions. We computed the COVID-19-related hospitalizations in the US considering four scenarios: four times the deaths (e.g., Italy), eight times the deaths (e.g., Hubei), ten times the deaths (e.g., close to current NY city) and 15 times the deaths (e.g., similar to Florida). We used then Monte Carlo simulations (1 million iterations) to compute the point estimate with 95% confidence intervals of number of infections among healthcare workers in the US if the hospitalizations would remain between four and eight times the deaths. These simulations were run for estimates based on Hubei and Italy separately. We adjusted our estimates for the number of hospital workers per bed. There are 5.06 healthcare workers per hospital bed in the US as compared to 3.36 in Italy and 1.08 in China.^24,25^ The adjustment was done by a factor of 1.50 for Italian estimates and 4.67 for Hubei estimates.

We then estimated the number of hospital workers infections if 100% of workers are provided PPE and PPEs are used properly by all hospital workers. For estimating infection counts if only the high risk (exposed) workers received PPE, we assumed that 30% of total healthcare would work directly with patients (i.e., high-risk) whereas 70% would be managing other patients as observed in China.^26^ For these estimations, we assumed the infection rate would be 55% in high risk (exposed) hospital workers and 26% in other hospital workers under inappropriate PPE conditions.^26^ We used the following to compute infections if PPE is available only in high risk workers.

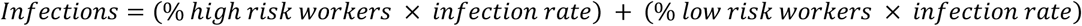

Based on age distribution of infection risks in Italy and China, we also computed COVID-19 infections if hospital workers over the age of 60 or 50 years are restricted from working directly with the patients.^5,27^ For all computations, we estimated the death rate assuming it to be 3% of infections as observed globally for cases upto March 27, 2020.^28^

## RESULTS

As of 3/19/2020, the province of Hubei had 67,800 cases of which 48,557 were from Wuhan and South Korea had 8,799 cases. As of 3/27/2020, Italy saw 79,968 infections. The mortality rate per million showed high variability between the regions (Table 1).

**Table 1:**
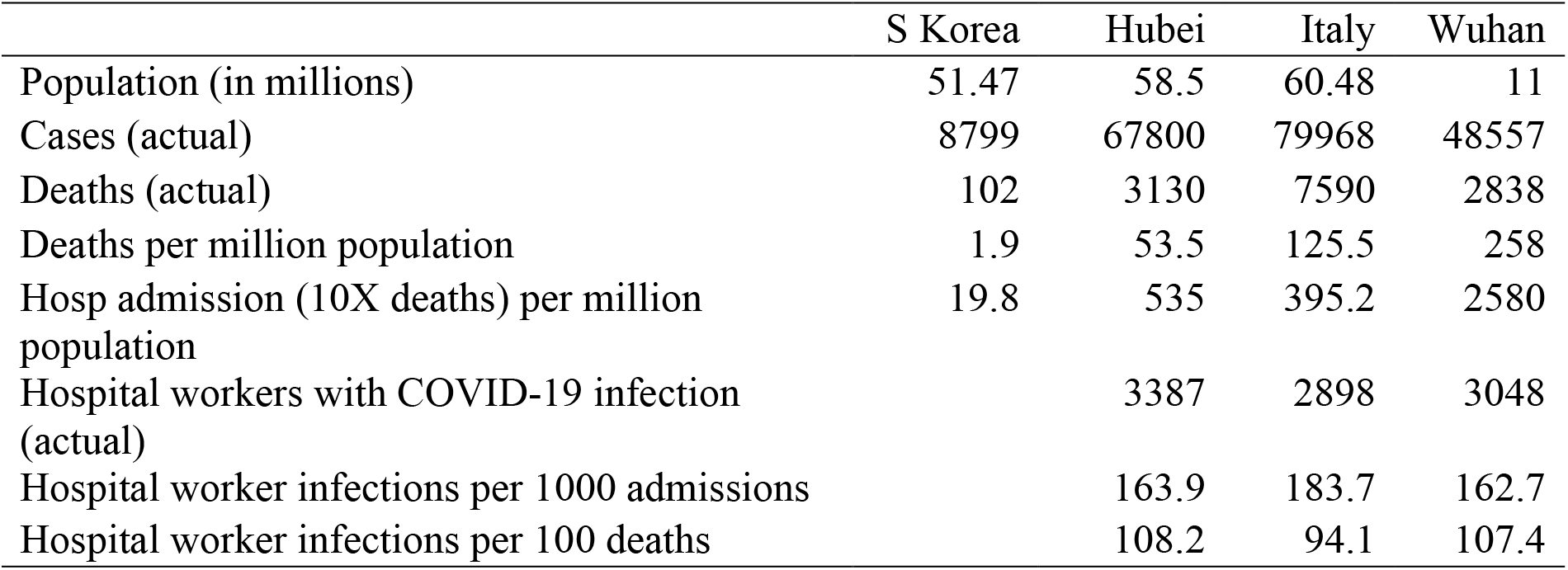
Summary of the total cases and infections and deaths amongst hospital workers in Hubei province, China and Italy

|  | S Korea | Hubei | Italy | Wuhan |
| --- | --- | --- | --- | --- |
| Population (in millions) | 51.47 | 58.5 | 60.48 | 11 |
| Cases (actual) | 8799 | 67800 | 79968 | 48557 |
| Deaths (actual) | 102 | 3130 | 7590 | 2838 |
| Deaths per million population | 1.9 | 53.5 | 125.5 | 258 |
| Hosp admission (10X deaths) per million population | 19.8 | 535 | 395.2 | 2580 |
| Hospital workers with COVID-19 infection (actual) |  | 3387 | 2898 | 3048 |
| Hospital worker infections per 1000 admissions |  | 163.9 | 183.7 | 162.7 |
| Hospital worker infections per 100 deaths |  | 108.2 | 94.1 | 107.4 |

Table 2 shows the projected numbers for death and hospital admission in the US. We used mortality rates from the four regions to estimate the total US mortality from worst (Wuhan) to the best (South Korea). We then calculated the expected number of patients based on the admission/death ratio of 4:1, 8:1, 10:1 and 15:1. If the epidemic in the US is as severe as in Wuhan with a similar health system response, then total admission may range from 196,383 to about 738,141 patients. Based on Hubei scenario, the US could see COVID-19 related hospitalizations from 54,996 to 206,236. Based on Italian scenario, the COVID-19 admissions in US could range from 165,379 to 620,170.

**Table 2.** Projected mortality and hospital admissions in United States given statistics from S. Korea, Hubei, Italy, and Wuhan

|  | S Korea<br>scenario | Hubei<br>scenario | Italy<br>scenario | Wuhan<br>scenario |
| --- | --- | --- | --- | --- |
| Projected total mortality in the US | 652 | 13749 | 41355 | 84998 |
| Admission projections (4X mortality) | 2609 | 54996 | 165379 | 196838 |
| Admission projections (8X mortality) | 5218 | 109992 | 330757 | 393675 |
| Admission projections (10X mortality) | 6523 | 137490 | 413447 | 492094 |
| Admission projections (15X mortality) | 9785 | 206236 | 620170 | 738141 |

Table 3 presents the unadjusted counts for hospital worker infections and deaths for scenarios if admissions were four, eight, ten or fifteen times the deaths. The highest number assumes a very high intensity epidemic with a large number of deaths and the health system is not prepared to handle the surge.

**Table 3.** Unadjusted estimated counts of COVID-19 infections in US hospital workers

|  | Hubei Model | Italy Model |
| --- | --- | --- |
| Admissions 4X of deaths |  |  |
| Infections | 9017 | 23588 |
| Admissions 8X of deaths |  |  |
| Infections | 18034 | 47177 |
| Admissions 10X of deaths |  |  |
| Infections | 22542 | 58971 |
| Admissions 15X of deaths |  |  |
| Infections | 33814 | 88456 |

The Monte Carlo simulations based on Hubei (China) suggested that about 53,640 US hospital workers (95% CI: 43,160 to 62,251) might get infected with COVID-19 after adjusting for differences between US and Chinese workers per beds. Similarly, the Monte Carlo simulations based on Italian estimates suggested that 53,097 US hospital workers (95% CI: 37,133 to 69,003) might get infected with COVID-19 after adjusting for differences between US and Italian workers per beds.

The detailed estimates for hospital worker infections and deaths under different scenarios are presented in Table 4. These estimates suggest that if ideal PPE conditions are implemented only for high-risk healthcare workers then the US might face 28,100 hospital worker infections (95% CI: 23,048 to 33,242) considering the Hubei (Chinese) scenario and about 28,354 hospital worker infections (95% CI: 19,829 to 36,848) considering the Italian scenario. Similarly, if hospital workers aged ≥ 60 years are restricted from direct patient care, then the US might face 1,985 hospital worker infections (95% CI: 1,627 to 2,347) based on the Hubei scenario and about 2,002 hospital worker infections (95% CI: 1,400 to 2,602) based on the Italian scenario.

**Table 4.** Adjusted estimated infection and death counts among hospital workers in US

|  | Hubei |  | Italy |  |
| --- | --- | --- | --- | --- |
|  | n<sub>e</sub> | 95% CI | n<sub>e</sub> | 95% CI |
| Infections (Total estimated) | 53640 | 43160-62251 | 53097 | 37133-69003 |
| 100% PPE for all hospital workers | 0* |  | 0* |  |
| 100% PPE for high-risk workers | 28100 | 23048-33242 | 28354 | 19829-36848 |
| Restricting workers age to < 60y | 1985 | 1627-2347 | 2002 | 1400-2602 |
| Restricting workers age to workers < 50y | 564 | 462-667 | 569 | 398-739 |
| Deaths (Total estimated) | 1579 | 1295-1868 | 1592 | 1114-2070 |
| 100% PPE for all hospital workers | 0* |  | 0* |  |
| 100% PPE for high-risk workers | 843 | 691-997 | 851 | 595-1105 |
| Restricting workers age to < 60y | 60 | 49-70 | 60 | 42-78 |
| Restricting workers age to workers < 50y | 17 | 14-20 | 17 | 12-22 |

n<sub>e</sub> n estimated
95% CI 95% Confidence Intervals
PPE Personal Protective Equipment
\* We assume that under ideal PPE conditions, there would be no infections and deaths among hospital care workers

Our analyses showed that death counts in US hospital workers could be as high as 1,579 (95% CI: 1,294 to 1867) based on Hubei scenario and could be around 1,592 (95% CI: 1,114 to 2,070) based on Italian scenario. The mortality estimates if the PPEs are only implemented for high-risk workers are 843 (95% CI: 691 to 997) under Hubei scenario and 851 (95% CI: 595 to 1105) under Italian scenario. The restriction of hospital workers aged ≥ 60 years from direct patient care would reduce the death counts to 60 based on the Hubei (95% CI: 49 to 70) and Italian (95% CI: 42 to 78) scenarios.

## DISCUSSION

We present estimates for the burden of disease and deaths due to COVID amongst hospital workers in the US. We highlight the risks of limited access or use of PPEs amongst hospital workers in the US and estimate the impact of two interventions: infection control including the use of PPEs and age restriction of hospital-based healthcare workers. We also present a clear path to significantly reducing this burden through two strategies: continuous wide-spread and proper use of personal protection strategies and limiting the exposure to hospital workers over the age of 60 years.

Unlike the community’s risk of exposure, we based the risk of hospital worker infections on the magnitude of exposure defined as the number of patients with COVID-19 admitted to the hospitals across the US. We believe that hospital admissions and deaths, rather than total number of cases in the community, are more useful measures of the risks to hospital workers and are key metrics of the burden on the healthcare system.

We based our estimates on several assumptions. Some of these assumptions are changing quite rapidly based on newer data. Our analysis assumes that the level of exposure of US hospital workers is similar to Italy and China. In both of these settings, especially in the earlier phase of the epidemic, there was a shortage of PPE which is also an issue in many US healthcare settings. Age is one of the key determinants of risk of infections and death in China and Italy.^14,15,29,30^ Our estimates of hospital admission are based on Italian and Chinese experience with different population distribution. In the US, earlier results show a relatively younger population group being affected.^29^ Similarly, the number of healthcare workers per bed is quite different in China, Italy and the US and was controlled in our analysis. We also controlled for the age distribution of the healthcare workforce including the age distribution of nurses and physicians in the US. We did not find data on other healthcare workers and assumed that their age distribution would be similar to nurses and physicians. We also assumed that the mortality rate amongst hospital will be three percent though the estimates for population level mortality have varied between 1.9 per million in South Korea to 258 per million to 395 per million in Lombardi, Italy. We were not able to do a sub-analysis of the mortality risk based on the presence of prevalent comorbidities like diabetes, heart diseases, and hypertension. We did not include some other interventions such as the shift length and number of hours working in the clinical areas. Earlier studies have found them to be a significant determinant of COVID-19 infection.^26^ Finally, we did not look at the mental health issues and the morbidity caused by them.

Infection control comprises multiple strategies such as those that focus on patients and patient location, healthcare providers with physical barriers and hygiene practices and the design of the healthcare facilities. We projected 100% effectiveness of infection control measures including PPEs based on studies conducted by Cheng et al in Hongkong and Schwartz et al from Taiwan during the SARS response.^31,32^ The PPE use and general infection control measures in both these studies were more liberal than the current recommendations by the CDC.^4^ For example, N95 respirators were recommended for taking care of all patients with suspected or confirmed COVID-19 and surgical masks were used in areas with no direct patient contact. Besides PPE, the intervention in these studies included hand hygiene compliance assessments, staff training and discussion as well as early testing with a turnaround time of 4-8 hours and patient isolation. There is a significant worry about the shortages of PPEs worldwide. Our study shows the potential impact of PPEs and other aggressive infection control measures Importantly, our study also shows that aggressive infection control measures including the use PPEs by all healthcare workers and not just high-risk workers should be considered. Implementation of such strategies will have to be balanced against the potential shortages in the high risk patient care areas, increased cost of healthcare delivery, decrease in the efficiency of care delivery, and in some cases potential for increase in the risk of infection if proper donning and doffing procedures are not adopted.

Age is the major determinant of death as in the general population including healthcare workers. 3,14,26 Based on the mortality data from Italy and China, we believe that reducing exposure of hospital workers over 60 could result in significant reduction in the overall morbidity and mortality amongst hospital workers.^14,26^ There is concern in the US that the age distribution is potentially different.^29^ If the age distribution is in fact younger, then an earlier age cut-off point can be considered. Currently, in the US, almost 30% of licensed physicians and nurses are over the age of 60 years and the impact of removing that large a workforce could be immense.^33-35^ Innovative solutions, such as the use of telemedicine, could limit the exposure of over 60 hospital workers while ensuring access to care to healthcare needs of the population.^36^

## Conclusion

We estimate a significant burden of illness and deaths due to COVID-19 if no strategies are adopted. We propose widespread availability and training on the use of PPEs to not only those working in highs risk areas of the hospital but to everyone providing direct patient care can significantly reduce the number of infected hospital workers. Similarly, reducing exposure of hospital workers above the age of 60 years, will reduce the death rates by over 90% amongst hospital workers but would require solutions that ensure service delivery to patients.

## Data Availability

The data for this study is available from corresponding author.

## DISCLAIMERS

The opinions and findings presented in this manuscript are of authors only. The views presented here do not represent the official position or policy of the authors’ affiliate institutions.

## CONTRIBUTION STATEMENT

JAR and OPR conceived they study and wrote the first draft. JAR, MRT and JAB performed the analyses and supported the draft of the study. All authors read the final version of results and manuscript.

## ETHICS AND REPORTING

The study uses the open source secondary anonymized data sources. The study meets the STROBE guidelines for reporting.

## REFERENCES

1. World Health Organization (WHO). Coronavirus disease 2019 (COVID-19) Situation Report – 61. Geneva: WHO; 2020.

2. Dong E, Du H, Gardner L. An interactive web-based dashboard to track COVID-19 in real time. Lancet Infect Dis. 2020.

3. Remuzzi A, Remuzzi G. COVID-19 and Italy: what next? Lancet. 2020.

4. Centers for Disease Control and Prevention (CDC). Interim Infection Prevention and Control Recommendations for Patients with Suspected or Confirmed Coronavirus Disease 2019 (COVID-19) in Healthcare Settings. Atlanta, GA: CDC;2020. [Available at URL: https://www.cdc.gov/coronavirus/2019-ncov/infection-control/control-recommendations.html] [Last Accessed: 03/24/2020].

5. Istituto Superiore di Sanità. Integrated surveillance of COVID-19 in Italy. Rome: Istituto Superiore di Sanità;2020. [Available at URL: https://www.epicentro.iss.it/coronavirus/bollettino/Infografica_27marzo%20ENG.pdf] [Last Accessed: 03/27/2020].

6. Rimmer A. Covid-19: GPs call for same personal protective equipment as hospital doctors. BMJ. 2020;368:m1055.

7. American College of Emergency Physicians (ACEP). COVID-19 Personal Protective Equipment (PPE) During the Pandemic. In. Irving, TX: ACEP; 2020 [Available at URL: https://www.acep.org/patient-care/policy-statements/covid-19-personal-protective-equipment-ppe-during-the-pandemic/] [Last Accessed: 03/24/2020].

8. Livingston E, Desai A, Berkwits M. Sourcing personal protective equipment during the COVID-19 pandemic. JAMA. 2020 Mar 28: doi:10.1001/jama.2020.5317.

9. Wong J, Goh QY, Tan Z, et al. Preparing for a COVID-19 pandemic: a review of operating room outbreak response measures in a large tertiary hospital in Singapore. Can J Anaesth. 2020.

10. Ng K, Poon BH, Kiat Puar TH, et al. COVID-19 and the Risk to Health Care Workers: A Case Report. Ann Intern Med. 2020.

11. Lu KL, Chen S, Leung LP. Initial Experience of an Emergency Department in Shenzhen in Responding to the Emerging Wuhan Coronavirus Pneumonia. Ann Emerg Med. 2020;75(4):556.

12. Gudi SK, Tiwari KK. Preparedness and Lessons Learned from the Novel Coronavirus Disease. Int J Occup Environ Med. 2020;11(2):108–112.

13. Madjid M, Safavi-Naeini P, Solomon SD, Vardeny O. Potential Effects of Coronaviruses on the Cardiovascular System: A Review. JAMA Cardiol. 2020.

14. Sun Y, Koh V, Marimuthu K, et al. Epidemiological and Clinical Predictors of COVID-19. Clin Infect Dis. 2020.

15. Fan J, Liu X, Pan W, Douglas MW, Bao S. Epidemiology of 2019 Novel Coronavirus Disease-19 in Gansu Province, China, 2020. Emerg Infect Dis. 2020;26(6).

16. Grasselli G, Pesenti A, Cecconi M. Critical Care Utilization for the COVID-19 Outbreak in Lombardy, Italy: Early Experience and Forecast During an Emergency Response. JAMA. 2020.

17. COVID-19 National Emergency Response Center ECMT, K.rea Centers for Disease Control & Prevention. Contact Transmission of COVID-19 in South Korea: Novel Investigation Techniques for Tracing Contacts. Osong Public Health Res Perspect. 2020;11(1):60–63.

18. Shim E, Tariq A, Choi W, Lee Y, Chowell G. Transmission potential and severity of COVID-19 in South Korea. Int J Infect Dis. 2020.

19. Zastrow M. South Korea is reporting intimate details of COVID-19 cases: has it helped? Nature. 2020.

20. DXY. COVID-19 Global Pandemic Real-Time Report. Beijing: DXY;2020. [Available at URL: http://ncov.dxy.cn/ncovh5/view/en_pneumonia?from=dxy&source=dxys] [Last Accessed: 03/30/2020] [Online].

21. New York City Department of Health. Coronavirus Disease 2019 (COVID-19). New York, NY: New York City Department of Health; 2020. [Available at URL: https://www1.nyc.gov/site/doh/covid/covid-19-main.page] [Last accessed: 03/30/2020].

22. Massachusetts Department of Public Health. Coronavirus Disease 2019 (COVID-19) Cases in MA. In. Boston, MA: Massachusetts Department of Public Health; 2020.

23. Florida Department of Health. Coronavirus: summary of persons being monitored, persons under investigation, and cases. Tallahassee, FL: Florida Department of Health;2020. [Available at URL: https://floridadisaster.org/globalassets/covid-19-data---daily-report-2020-03-26-1823.pdf] [Last accessed: 03/28/2020].

24. Organisation for Economic Co-operation and Development (OECD). OECD Health Statistics 2019. In. Paris: OECD; 2019.

25. American Hospital Association. Fast Facts on US Hospitals. Chicago, IL: American Hospital Association; 2020. [Available at URL: https://www.aha.org/system/files/media/file/2020/01/2020-aha-hospital-fast-facts-new-Jan-2020.pdf] [Last accessed: 03/30/2020].

26. Ran L, Chen X, Wang Y, Wu W, Zhang L, Tan X. Risk Factors of Healthcare Workers with Corona Virus Disease 2019: A Retrospective Cohort Study in a Designated Hospital of Wuhan in China. Clin Infect Dis. 2020.

27. Liu R, Han H, Liu F, et al. Positive rate of RT-PCR detection of SARS-CoV-2 infection in 4880 cases from one hospital in Wuhan, China, from Jan to Feb 2020. Clin Chim Acta. 2020;505:172–175.

28. Wu JT, Leung K, Bushman M, et al. Estimating clinical severity of COVID-19 from the transmission dynamics in Wuhan, China. Nature Medicine. 2020. doi: 10.1038/s41591-020-0822-7.

29. Team CC-R. Severe Outcomes Among Patients with Coronavirus Disease 2019 (COVID-19) - United States, February 12-March 16, 2020. MMWR Morb Mortal Wkly Rep. 2020;69(12):343–346.

30. Khot WY, Nadkar MY. The 2019 Novel Coronavirus Outbreak - A Global Threat. J Assoc Physicians India. 2020;68(3):67–71.

31. Cheng VCC, Wong SC, Chen JHK, et al. Escalating infection control response to the rapidly evolving epidemiology of the Coronavirus disease 2019 (COVID-19) due to SARS-CoV-2 in Hong Kong. Infect Control Hosp Epidemiol. 2020:1–24.

32. Schwartz J, King CC, Yen MY. Protecting Health Care Workers during the COVID-19 Coronavirus Outbreak -Lessons from Taiwan’s SARS response. Clin Infect Dis. 2020.

33. Zhang X, Lin D, Pforsich H, Lin VW. Physician workforce in the United States of America: forecasting nationwide shortages. Hum Resour Health. 2020;18(1):8.

34. Juraschek SP, Zhang X, Ranganathan V, Lin VW. United States Registered Nurse Workforce Report Card and Shortage Forecast. Am J Med Qual. 2019;34(5):473–481.

35. Skinner L, Staiger DO, Auerbach DI, Buerhaus PI. Implications of an Aging Rural Physician Workforce. N Engl J Med. 2019;381(4):299–301.

36. Buerhaus PI, Auerbach DI, Staiger DO. Older Clinicians and the Surge in Novel Coronavirus Disease 2019 (COVID-19). JAMA. 2020.

